# Prior COVID-19 Infection and Antibody Response to Single Versus Double Dose mRNA SARS-CoV-2 Vaccination

**DOI:** 10.1101/2021.02.23.21252230

**Authors:** Joseph E. Ebinger, Justyna Fert-Bober, Ignat Printsev, Min Wu, Nancy Sun, Jane C. Figueiredo, Jennifer E. Van Eyk, Jonathan G. Braun, Susan Cheng, Kimia Sobhani

## Abstract

The double dose regimen for mRNA vaccines against SARS-CoV-2 presents both a hope and a challenge for global efforts to curb the COVID-19 pandemic. With supply chain logistics impacting the rollout of population-scale vaccination programs, increasing attention has turned to the potential efficacy of single versus double dose vaccine administration for select individuals. To this end, we examined response to Pfizer-BioNTech mRNA vaccine in a large cohort of healthcare workers including those with versus without prior COVID-19 infection. For all participants, we quantified circulating levels of SARS-CoV-2 anti-spike (S) protein IgG at baseline prior to vaccine, after vaccine dose 1, and after vaccine dose 2. We observed that the anti-S IgG antibody response following a single vaccine dose in persons who had recovered from confirmed prior COVID-19 infection was similar to the antibody response following two doses of vaccine in persons without prior infection (P≥0.58). Patterns were similar for the post-vaccine symptoms experienced by infection recovered persons following their first dose compared to the symptoms experienced by infection naïve persons following their second dose (P=0.66). These results support the premise that a single dose of mRNA vaccine could provoke in COVID-19 recovered individuals a level of immunity that is comparable to that seen in infection naïve persons following a double dose regimen. Additional studies are needed to validate our findings, which could allow for public health programs to expand the reach of population wide vaccination efforts.

## INTRODUCTION

Rapid development of vaccines against SARS-CoV-2, the causative agent of Coronavirus Disease 2019 (COVID-19), offers great promise for curbing spread of infection and accelerating the timeline towards a potential level of herd immunity.^1-3^ Amidst ongoing efforts to rapidly deploy vaccinations, challenges to the supply chain have prompted queries around whether single rather than double dose administration may suffice for certain individuals – including those recovered from prior COVID-19 infection.^4^ Emerging data from small studies suggest that individuals who have recovered from either a recent or remote COVID-19 infection may have a sustained immunity that could be assessed via measurable antibody response to a single vaccine dose administration.^5,6^ To this end, we evaluated the SARS-CoV-2 antibody response following first and second doses of mRNA vaccination administered in a large and diverse cohort of healthcare workers while specifically focusing on the response in persons with confirmed prior COVID-19 compared to those without prior infection.

## METHODS

In a cohort of healthcare workers who received Pfizer-BioNTech vaccination at our medical center in Southern California,^7^ we used the Abbott Architect immunoassays (Abbott Park, IL) to quantify circulating levels of SARS-CoV-2 anti-nucleocapsid (N) protein IgG and anti-spike (S) protein IgG at 3 time points: before or up to 3 days after dose 1, within 7 to 21 days after dose 1, and within 7 to 21 days after dose 2. The Abbott anti-S IgG assay is CE marked with anticipated near-future emergency use authorization. Given that a conservative high titer plaque reduction neutralization (PRNT) assay of 1:250 has been correlated to the anti-S IgG cutoff of 4160 AU/mL, we additionally examined the proportion of vaccine recipients who achieved this threshold following administration of one dose or two.

All participants provided survey data on prior COVID-19 infection and symptoms experienced after each vaccine dose. We determined prior infection status and timing in relation to first vaccine date, based on concordance of data documented in health records, presence of anti-N IgG antibodies at baseline pre-vaccination testing, and the self-reported survey information collected. All cases of data discrepancy regarding prior COVID-19 infection status underwent manual physician adjudication.

We compared antibody level and symptom responses between those with and without a prior COVID-19 diagnosis. We analyzed data at both matched and shifted time points, including examining measures for those with a prior COVID-19 diagnosis (at baseline and following dose 1) compared to those without a prior COVID-19 diagnosis (following dose 1 and dose 2). We log-transformed non-normally distributed values. For comparing between-group continuous values, we used the non-parametric Wilcoxon Rank Sum test. For comparing between-group proportions of values above a give threshold, we used two-sided Chi-square tests. We performed sensitivity analyses for those participants with immunoassays at all 3 time points. We conducted all statistical analyses using R (v3.6.1) and considered statistical significance as a two-tailed P value <0.05. All participants provided written informed consent; all protocols were approved by the Cedars-Sinai institutional review board.

## RESULTS

A total of N=1089 vaccine recipients provided at least one blood sample for antibody testing, with an average age of 41.8±12.1 years, 60.8% female and 53.4% non-white (**Table 1**). Of this total sample, N=980 (78 with prior COVID-19 infection) provided baseline (pre-vaccine) samples, N=525 (35 with prior infection) provided samples after dose 1, and 238 (11 with prior infection) provided samples after dose 2. A total of 216 individuals (10 with prior infection) provided blood samples at all 3 time points.

**Table 1.**
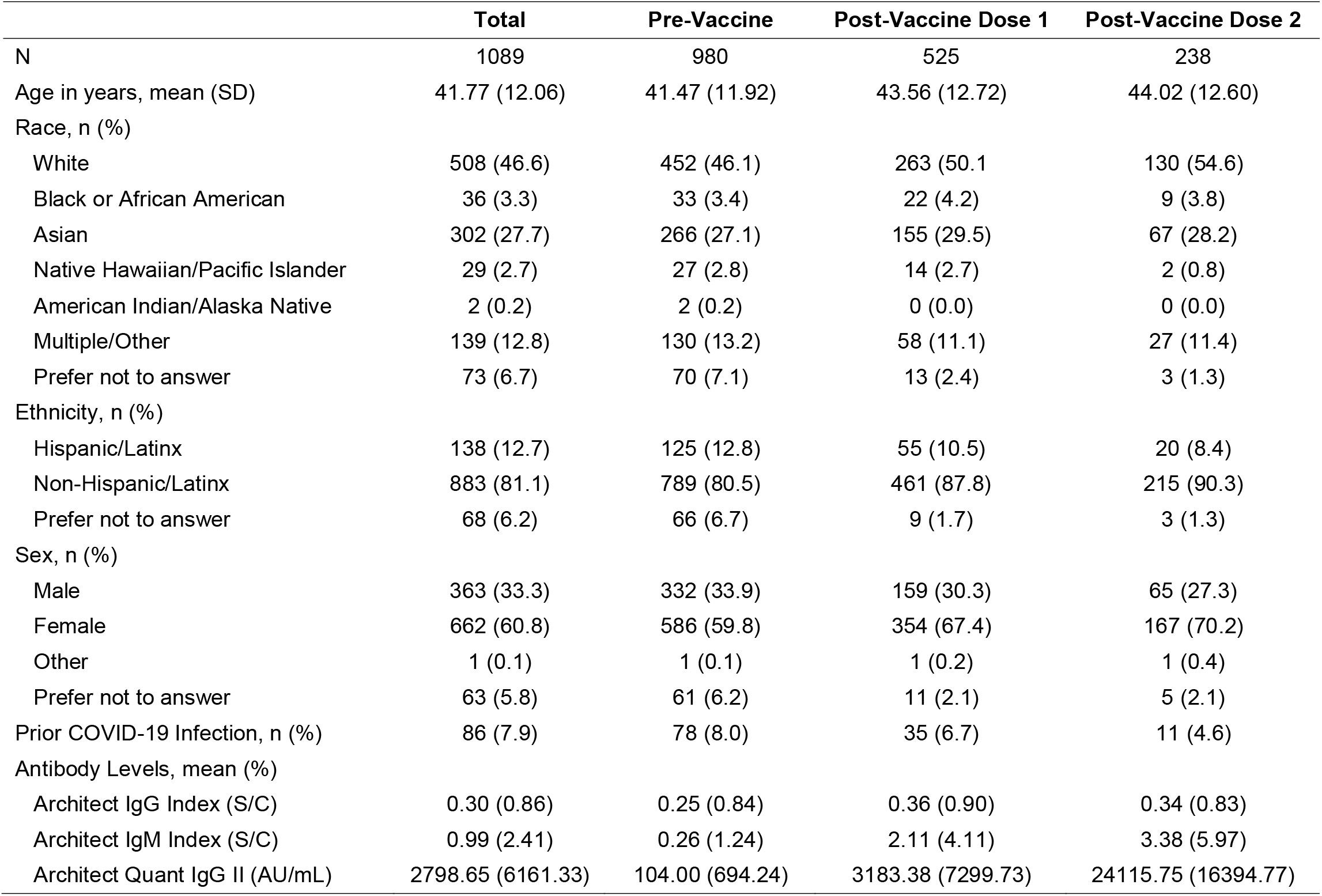
Characteristics of the Study Sample.

|  | **Total** | **Pre-Vaccine** | **Post-Vaccine Dose 1** | **Post-Vaccine Dose 2** |
| --- | --- | --- | --- | --- |
| N | 1089 | 980 | 525 | 238 |
| Age in years, mean (SD) | 41.77 (12.06) | 41.47 (11.92) | 43.56 (12.72) | 44.02 (12.60) |
| Race, n (%) |  |  |  |  |
| White | 508 (46.6) | 452 (46.1) | 263 (50.1) | 130 (54.6) |
| Black or African American | 36 (3.3) | 33 (3.4) | 22 (4.2) | 9 (3.8) |
| Asian | 302 (27.7) | 266 (27.1) | 155 (29.5) | 67 (28.2) |
| Native Hawaiian/Pacific Islander | 29 (2.7) | 27 (2.8) | 14 (2.7) | 2 (0.8) |
| American Indian/Alaska Native | 2 (0.2) | 2 (0.2) | 0 (0.0) | 0 (0.0) |
| Multiple/Other | 139 (12.8) | 130 (13.2) | 58 (11.1) | 27 (11.4) |
| Prefer not to answer | 73 (6.7) | 70 (7.1) | 13 (2.4) | 3 (1.3) |
| Ethnicity, n (%) |  |  |  |  |
| Hispanic/Latinx | 138 (12.7) | 125 (12.8) | 55 (10.5) | 20 (8.4) |
| Non-Hispanic/Latinx | 883 (81.1) | 789 (80.5) | 461 (87.8) | 215 (90.3) |
| Prefer not to answer | 68 (6.2) | 66 (6.7) | 9 (1.7) | 3 (1.3) |
| Sex, n (%) |  |  |  |  |
| Male | 363 (33.3) | 332 (33.9) | 159 (30.3) | 65 (27.3) |
| Female | 662 (60.8) | 586 (59.8) | 354 (67.4) | 167 (70.2) |
| Other | 1 (0.1) | 1 (0.1) | 1 (0.2) | 1 (0.4) |
| Prefer not to answer | 63 (5.8) | 61 (6.2) | 11 (2.1) | 5 (2.1) |
| Prior COVID-19 Infection, n (%) | 86 (7.9) | 78 (8.0) | 35 (6.7) | 11 (4.6) |
| Antibody Levels, mean (%) |  |  |  |  |
| Architect IgG Index (S/C) | 0.30 (0.86) | 0.25 (0.84) | 0.36 (0.90) | 0.34 (0.83) |
| Architect IgM Index (S/C) | 0.99 (2.41) | 0.26 (1.24) | 2.11 (4.11) | 3.38 (5.97) |
| Architect Quant IgG II (AU/mL) | 2798.65 (6161.33) | 104.00 (694.24) | 3183.38 (7299.73) | 24115.75 (16394.77) |

For both anti-N IgG (representing response to prior infection) and anti-S IgG (representing response to either prior infection or vaccine), COVID-19 recovered persons had expectedly higher antibody levels at all time points (P≤0.001) (**Tables S1-S2** and **Figures S1-S2**). Notably, anti-S IgG levels were only slightly lower in COVID-19 recovered persons at baseline, when compared to infection naïve persons who had received a single vaccine dose (log median AU/mL [IQR], 6.0 [4.6, 6.9] vs 7.0 [6.3, 7.6], P<0.001). Moreover, anti-S IgG levels were not significantly different between COVID-19 recovered persons following a single dose and infection naïve persons who had received 2 doses (10.0 [9.2, 10.4] vs 9.9 [9.4, 10.3], P=0.91) (**Figure 1**). In parallel with analyses of antibody response, we also observed that overall frequency and severity of post-vaccine symptoms were more prominent for COVID-19 recovered after dose 1 compared to infection naïve persons after dose 2 (**Figure S3**).

**Figure 1.**
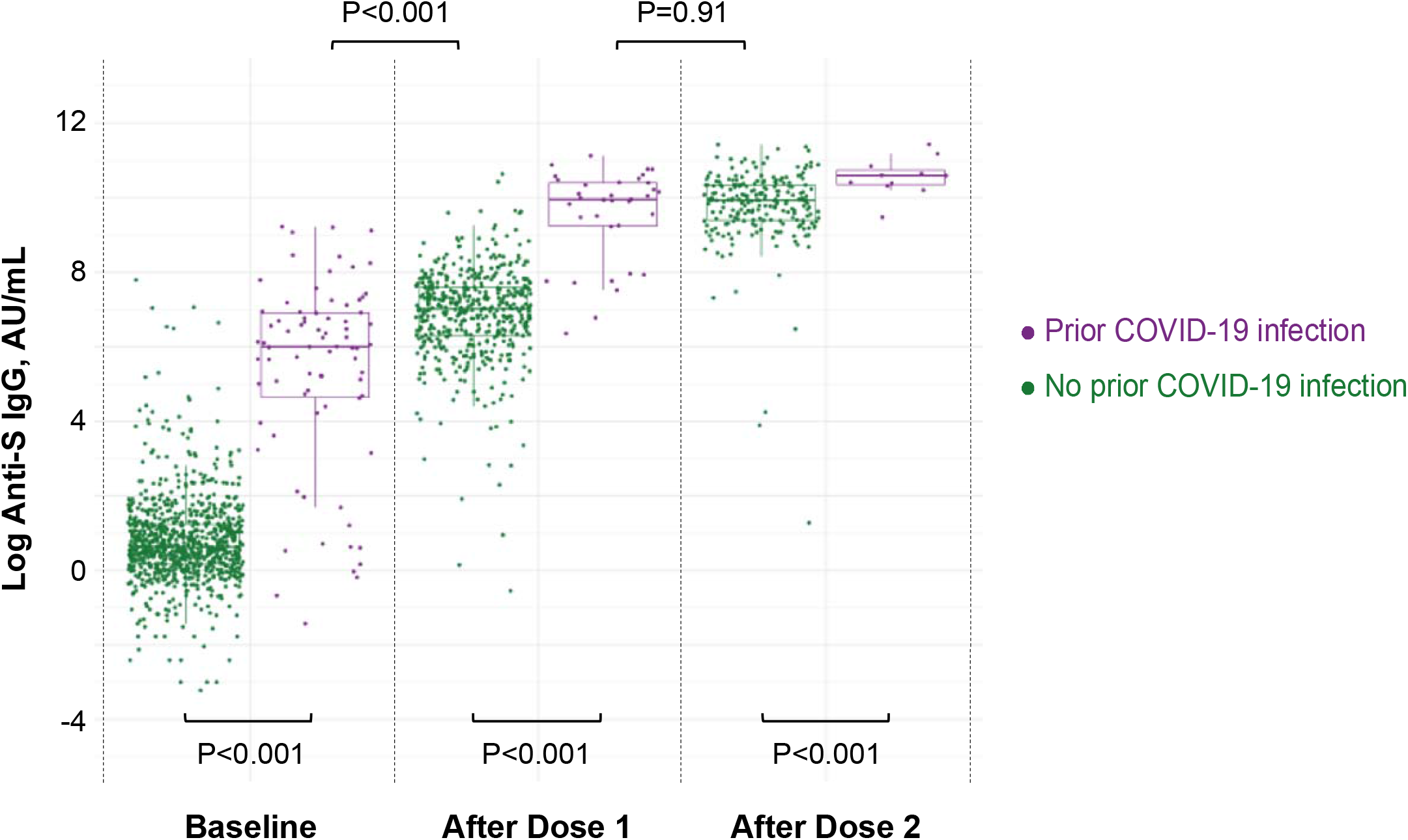
Anti-Spike IgG Antibody Response to mRNA SARS-CoV-2 Vaccination in Persons With and Without Prior COVID-19 Infection. Box plots display the median values with the interquartile range (lower and upper hinge) and ±1.5-fold the interquartile range from the first and third quartile (lower and upper whiskers).

Similar results were found in a sensitivity analysis including only individuals who had antibody immunoassays performed at all 3 time points (**Tables S3-S4**). Specifically, those with prior COVID-19 infection had higher anti-S IgG than those without prior infection at all time points. No difference in anti-S IgG level was detected between those with a prior COVID-19 infection after one dose of vaccine compared with those without prior infection following two doses (10.2 [8.4, 10.5] vs 9.9 [9.4, 10.3], P=0.58). In sensitivity analyses, frequency and severity of post-vaccine symptoms were also similar to those seen in the main analyses.

In the total sample, proportions of anti-S IgG levels at or above the 4160 AU/mL threshold were similar in COVID-19 recovered persons at baseline compared to infection naïve persons after a single dose (P=0.94). Notably, these proportions were lower in COVID recovered after a single dose compared to infection naïve persons after 2 doses (P<0.001), and then no different when both groups were compared after 2 doses (**Table S5** and **Figure S4**).

## DISCUSSION

We found that COVID-19 recovered individuals develop a level of provoked antibody response, following a single dose of mRNA vaccine, that is comparable to the provoked antibody response seen after a two-dose vaccination course administered to infection naïve persons. Extending from similar results seen in smaller studies,^5,6^ our findings in a large and diverse cohort of healthcare workers highlight the potential of a strategy for maximizing vaccine supply that warrants further investigation.

Recent work has demonstrated that COVID-19 specific antibodies are efficiently generated and detectable in the circulation following a single dose of vaccines that were originally intended for complete administration to include an additional booster dose.^5^ These findings have prompted some organizations to favor prioritizing at least a first vaccine dose to majority segments of the population while considering variable timing for the second dose. In the absence of clinical outcomes data to support any variations from pre-specified vaccination protocols,^8-10^ there are immuno-biological data suggesting possible alternate strategies for COVID-19 recovered individuals. In fact, detectable presence of naturally acquired anti-SARS-CoV-2 antibodies and measures of discernible T-cell mediated immunity, especially in persons who have successfully recovered from recent versus remote infection, have prompted some experts to suggest delaying any vaccination for these individuals.^11-13^ However, acknowledging the unclear duration of naturally acquired immunity and the unknown extent to which immunity to one strain of SARS-CoV-2 confers protection from variants, there is general agreement that vaccination strategies for COVID-19 recovered persons warrants careful consideration.

Our data suggest the potential benefit of at least one vaccine dose, given that we observed pre-vaccine levels of anti-S IgG in COVID-19 recovered persons to be somewhat lower than levels detected among infection naïve persons following a single vaccine dose. In the absence of directly measured neutralizing antibody levels, we secondarily examined the proportion of anti-S IgG values known to correspond with high titers of the PRNT assay and found a comparable albeit slightly lower frequency in COVID-19 recovered persons after a single vaccine dose compared to infection naïve persons after two doses. Notwithstanding the need for further studies with additional serologic phenotyping, this finding may be related to heterogeneity within the COVID-19 recovered persons including variations in timing and severity of prior illness. Although circulating antibody levels alone are not definitive measures of immune status, serial measures of the serological response to either natural or inoculated exposures are known to correlate well with effective immunity^14^ and our results indicate their potential utility in guiding vaccine deployment strategies for both infection recovered and naïve persons.

Several limitations of our study merit consideration. We examined antibody response within 7 to 21 days following each vaccine dose, and longer-term follow up is likely to provide additionally informative data – particular regarding the putative duration of immunity acquired from receiving a single versus double dose of vaccine administration. Direct measures of neutralizing antibody levels are a part of our ongoing work. Notwithstanding the size of our study cohort, yet larger-sized samples are needed for sufficient statistical power to examine differences across demographic and clinical subgroups that are known to exhibit variation in antibody response following vaccination.^15-17^

In summary, we found in diverse cohort of mRNA vaccine recipients that anti-S IgG levels are similar between those with and without prior COVID-19 infection after receiving their first and second doses, respectively. These results offer preliminary evidence in support of a middle ground between public health motivated and immunologically supported vaccine strategies. If validated, an approach that involves providing a single dose of vaccine to persons with a confirmed history of COVID-19 infection along with an on-time complete vaccine schedule for infection naïve persons could assist with maximizing the benefit of a limited vaccine supply.

## Data Availability

The data that support the findings of this study are available from Cedars-Sinai Medical Center, upon reasonable request. The data are not publicly available due to the contents including information that could compromise research participant privacy/consent.

## ACKNOWLEDGEMENTS

We are grateful to all the front-line healthcare workers in our healthcare system who continue to be dedicated to delivering the highest quality care for all patients. We would like to thank the following people for their collective effort: Francesca Paola Aguirre, MD; Kawsar Ahmad; Christine M. Albert, MD, MPH; Mona Alotaibi, MD; Allen Andres, PhD; Moshe Arditi, MD; Ani Balmanoukian, MD; Courtney Becker; James Beekley, MPH; Diana Benliyan; Anders H. Berg, MD, PhD; Eva Biener-Ramanujan; Aleksandra Binek, PhD; Patrick Botting, MSPH; Gregory J. Botwin, BS; David Casero; Cindy Chavira, MBA; Blandine Chazarin Orgel; Mingtian Che; Peter Chen, MD; Vi Chiu, MD, PhD; Dain Choi; Melanie Chow; Cathie Chung; MD; Cailin Climer; Bernice Coleman, PhD, RN; Sandra Contreras, MPH; Rachel Coren, MPH; Donna Costales, RN; Wendy Cozen, DO, MPH; Tahir Dar; Jennifer Davis; Tod Davis; Philip Debbas; Jacqueline Diaz; Jessica Dos Santos, Matthew Driver, MPH; Keren R. Dunn, CIP; Rebecca Ely, RN; Mark Faries; Justyna Fert-Bober, PhD, MD; Barbara Fields, RN; Lucia Florindez, PhD; Joslyn Foley; Norma Fontelera; Sarah Francis, RN; Jeffrey A. Golden, MD; Alma Gonzalez; Helen S. Goodridge, PhD; Jeanette Gonzalez; Jonathan D. Grein, MD; Gena Guidry, MSc; Omid Hamid, MD; Mary Hanna; Shima Hashemzadeh; Mallory Heath, MLS; Ergueen Herrera; Amy Hoang, MS; Lilith Huang; Khalil Huballa; Quyen Hurlburt, RN; Shehnaz K. Hussain, PhD; Carissa A. Huynh; Justina Ibrahim; Ugonna Ihenacho, MPH; Mohit Jain, MD, PhD; Harneet Jawanda, MD; Mary Jordan, Ashley Jose-Isip; Sandy Joung, MHDS; Michael Karin, PhD; Elizabeth H. Kim, MHDS; Linda Kim, PhD; Michelle M. Kittleson, MD, PhD; Edward Kowalewski; Catherine N. Le, MD; Nicole A. Leonard, JD, MBA; Yin Li; Yunxian Liu, PhD; John Lloyd; Eric Luong, MPH; Anzhelya Makaryan; Bhavya Malladi; Danica-Mae Manalo, David Marshall, DNP, JD; Angela McArdle; Dermot P.B. McGovern, MD, PhD; Inderjit Mehmi, MD; Darlene Mejia; Gil Y. Melmed, MD; Larry Mendez; Emebet Mengesha; Akil Merchant, MD; Noah Merin, MD, PhD; Kathrin S. Michelsen, PhD; Gail Milan, RN; Peggy B. Miles, MD; Jordan Miller; Margo Minissian, PhD; Romalisa Miranda-Peats, MPH; Seyedeh Elnaz Mirzadeh; April Moore; Pamela Moore; Janette Moreno, DNP; Angela Mujukian, MD; Nathalie Nguyen, MPH; Trevor Trung Nguyen; Magali Noval Rivas, PhD; Fleury Nsole Bitghe, PhD; Michelle Offner, NP; Jillian Oft, MD; Elmar Park; Eunice Park; Vipul Patel, PharmD; Connor Phebus; Lawrence Piro, MD; Lauren R. Polak, JD; Ashley Porter; Matthew Puccio; Koen Raedschelders, PhD; V. Krishnan Ramanujan, PhD; Rocio Ramirez; Gerardo Ramirez; Mohamad Rashid, MBChB; Karen Reckamp, MD; Kylie Rhoades; Celine E. Riera, PhD; Richard V. Riggs, MD; Alejandro Rivas; Jackie Robertson; Maria Salas; Michelle Schafieh, MS; Rita Shane, PharmD; Sonia Sharma, PhD; Cristina Simons, RN; Kimia Sobhani, PhD; Muhammad Soomar; Sarah Sternbach; Nancy Sun, MPS; Clive Svendsen, PhD; Brian Tep; Rose O. Tompkins, MD; Warren G. Toutellotte, MD, PhD; Rocio Vallejo; Christy Velasco; Mectabel Velasquez; Kirstin Washington; Kristopher Wentzel, MD; Shane White; Benjamin Wong; Melissa Wong, MD; Mahendra Yatawara, MBA; Rachel Zabner, MD; Cindy Zamudio, MD; Yi Zhang, MD, MS; Lisa Zhou; Patrick Zvara, MS.

## DATA AVAILABILITY

The data supporting the findings of this study are available from Cedars-Sinai Medical Center, upon reasonable request and via institutional data sharing policies. The data are not publicly available due to the contents containing information that could compromise participant privacy.

## CODE AVAILABILITY

The code used for all analyses is available upon reasonable request.

