## Supplemental Material for "Prior COVID-19 Infection and Antibody Response to Single Versus Double Dose mRNA SARS-CoV-2 Vaccination"

**Table S1. Comparison of antibody levels between participants with and without prior COVID-19 infection at matched time points.** Values are shown as median and interquartile range [IQR], with comparisons performed using Wilcoxon tests. Sample sizes at baseline, After Dose 1, and After Dose 2 included: N=902, N=490, and N=227 for persons without prior COVID-19; and N=78, N=35, and N=11 for persons with prior COVID-19.

| | No Prior COVID-19 | Prior COVID-19 | P value |
|---|---|---|---|
| **Log IgG(S)** | | | |
| Baseline | 0.6 [0.2, 1.2] | 6.0 [4.6, 6.9] | <0.001 |
| After Dose 1 | 7.0 [6.3, 7.6] | 10.0 [9.2, 10.4] | <0.001 |
| After Dose 2 | 9.9 [9.4, 10.3] | 10.6 [10.3, 10.7] | <0.001 |
| **Log IgG(N)** | | | |
| Baseline | -3.9 [-4.6, -3.2] | 0.6 [-0.5, 1.2] | <0.001 |
| After Dose 1 | -2.8 [-3.9, -1.7] | 0.6 [-0.1, 1.2] | <0.001 |
| After Dose 2 | -2.7 [-3.7, -1.3] | 1.0 [0.2, 1.1] | <0.001 |
| **Log IgM(S)** | | | |
| Baseline | -3.2 [-3.5, -2.7] | -0.3 [-1.4, 0.8] | <0.001 |
| After Dose 1 | 0.1 [-0.8, 0.8] | 0.1 [-0.4, 1.0] | 0.43 |
| After Dose 2 | 0.7 [-0.1, 1.3] | -0.1 [-0.6, 1.4] | 0.59 |

**Table S2. Comparison of antibody levels between participants with and without prior COVID-19 infection at shifted time points.** Values are shown as median and interquartile range [IQR], with comparisons performed using Wilcoxon tests. Sample sizes at baseline, After Dose 1, and After Dose 2 included: N=902, N=490, and N=227 for persons without prior COVID-19; and N=78, N=35, and N=11 for persons with prior COVID-19.

| | No Prior COVID-19 | Prior COVID-19 | P value |
|---|---|---|---|
| **No Prior COVID-19 After Dose 1, Prior COVID-19 at Baseline** | | | |
| Log IgG(S) | 7.0 [6.3, 7.6] | 6.0 [4.6, 6.9] | <0.001 |
| Log IgG(N) | -2.8 [-3.9, -1.7] | 0.6 [-0.5, 1.2] | <0.001 |
| Log IgM(S) | 0.1 [-0.8, 0.8] | -0.3 [-1.4, 0.8] | 0.09 |
| **No Prior COVID-19 After Dose 2, Prior COVID-19 After Dose 1** | | | |
| Log IgG(S) | 9.9 [9.4, 10.3] | 10.0 [9.2, 10.4] | 0.91 |
| Log IgG(N) | -2.7 [-3.7, -1.3] | 0.6 [-0.1, 1.2] | <0.001 |
| Log IgM(S) | 0.7 [-0.1, 1.3] | 0.1 [-0.4, 1.0] | 0.052 |

**Table S3. Sensitivity analysis comparing antibody levels between participants with and without prior COVID-19 infection at matched time points.** Values are shown as median and interquartile range [IQR], with comparisons performed using Wilcoxon tests. The sample with data available at all time points included N=206 without prior COVID-19 and N=10 with prior COVID-19.

| | No Prior COVID-19 | Prior COVID-19 | P value |
|---|---|---|---|
| **Log IgG(S)** | | | |
| Baseline | 0.7 [0.2, 1.2] | 5.9 [2.7, 7.2] | <0.001 |
| After Dose 1 | 7.0 [6.2, 7.6] | 10.2 [8.4, 10.5] | <0.001 |
| After Dose 2 | 9.9 [9.4, 10.3] | 10.6 [10.3, 10.8] | 0.001 |
| **Log IgG(N)** | | | |
| Baseline | -3.9 [-4.6, -3.0] | 0.7 [0.1, 1.0] | <0.001 |
| After Dose 1 | -2.7 [-3.9, -1.6] | 0.8 [-0.1, 1.2] | <0.001 |
| After Dose 2 | -2.7 [-3.9, -1.3] | 0.9 [0.0, 1.1] | <0.001 |
| **Log IgM(S)** | | | |
| Baseline | -3.2 [-3.5, -2.7] | -1.2 [-1.9, 0.5] | <0.001 |
| After Dose 1 | 0.0 [-0.7, 0.7] | 0.1 [-0.2, 1.0] | 0.51 |
| After Dose 2 | 0.7 [-0.1, 1.3] | -0.2 [-0.7, 1.1] | 0.24 |

**Table S4. Sensitivity analysis comparing antibody levels between participants with and without prior COVID-19 infection at shifted time points.** Values are shown as median and interquartile range [IQR], with comparisons performed using Wilcoxon tests. The sample with data available at all time points included N=206 without prior COVID-19 and N=10 with prior COVID-19.

| | No Prior COVID-19 | Prior COVID-19 | P value |
|---|---|---|---|
| **No Prior COVID-19 After Dose 1, Prior COVID-19 at Baseline** | | | |
| Log IgG(S) | 7.0 [6.2, 7.6] | 5.9 [2.7, 7.2] | 0.049 |
| Log IgG(N) | -2.7 [-3.9, -1.6] | 0.7 [0.1, 1.0] | 0.001 |
| Log IgM(S) | 0.0 [-0.7, 0.7] | -1.2 [-1.9, 0.5] | 0.08 |
| **No Prior COVID-19 After Dose 2, Prior COVID-19 After Dose 1** | | | |
| Log IgG(S) | 9.9 [9.4, 10.3] | 10.2 [8.4, 10.5] | 0.58 |
| Log IgG(N) | -2.7 [-3.9, -1.3] | 0.8 [-0.1, 1.2] | <0.001 |
| Log IgM(S) | 0.7 [-0.1, 1.3] | 0.1 [-0.2, 1.0] | 0.31 |

**Table S5. Comparison of proportions of anti-Spike protein IgG antibody levels ≥50 AU/mL between participants with and without prior COVID-19 infection.** Sample sizes at baseline, After Dose 1, and After Dose 2 included: N=902, N=490, and N=227 for persons without prior COVID-19; and N=78, N=35, and N=11 for persons with prior COVID-19.

| | No Prior COVID-19 | Prior COVID-19 | P value |
|---|---|---|---|
| **Matched time points** | | | |
| **IgG S ≥50 AU/mL** | **n/N (%)** | **n/N (%)** | |
| Baseline | 19/902 (2) | 62/78 (79) | <0.001 |
| After Dose 1 | 479/490 (98) | 35/35 (100) | 0.78 |
| After Dose 2 | 225/227 (99) | 11/11 (100) | 1.00 |
| **Shifted time points** | | | |
| ***No Prior COVID-19 After Dose 1, Prior COVID-19 at Baseline*** | | | |
| IgG(S) ≥50, % | 479/490 (98) | 62/78 (79) | <0.001 |
| ***No Prior COVID-19 After Dose 2, Prior COVID-19 After Dose 1*** | | | |
| IgG(S) ≥50, % | 225/227 (99) | 35/35 (100) | 1.00 |

**Table S6. Comparison of proportions of anti-Spike protein IgG antibody levels ≥4160 AU/mL between participants with and without prior COVID-19 infection.** Sample sizes at baseline, After Dose 1, and After Dose 2 included: N=902, N=490, and N=227 for persons without prior COVID-19; and N=78, N=35, and N=11 for persons with prior COVID-19.

| | No Prior COVID-19 | Prior COVID-19 | P value |
|---|---|---|---|
| **Matched time points** | | | |
| **IgG S ≥4160 AU/mL** | **n/N (%)** | **n/N (%)** | |
| Baseline | 0/902 (0) | 6/78 (8) | <0.001 |
| After Dose 1 | 37/490 (8) | 27/35 (77) | <0.001 |
| After Dose 2 | 220/227 (97) | 11/11 (100) | 1.00 |
| **Shifted time points** | | | |
| ***No Prior COVID-19 After Dose 1, Prior COVID-19 at Baseline*** | | | |
| IgG(S) ≥4160, % | 37/490 (8) | 6/78 (8) | 1.00 |
| ***No Prior COVID-19 After Dose 2, Prior COVID-19 After Dose 1*** | | | |
| IgG(S) ≥4160, % | 220/227 (97) | 27/35 (77) | <0.001 |

**Figure S1. Anti-Nucleocapsid IgG Antibody Response to mRNA SARS-CoV-2 Vaccination in Persons With and Without Prior COVID-19 Infection.** Box plots display the median values with the interquartile range (lower and upper hinge) and ±1.5-fold the interquartile range from the first and third quartile (lower and upper whiskers).

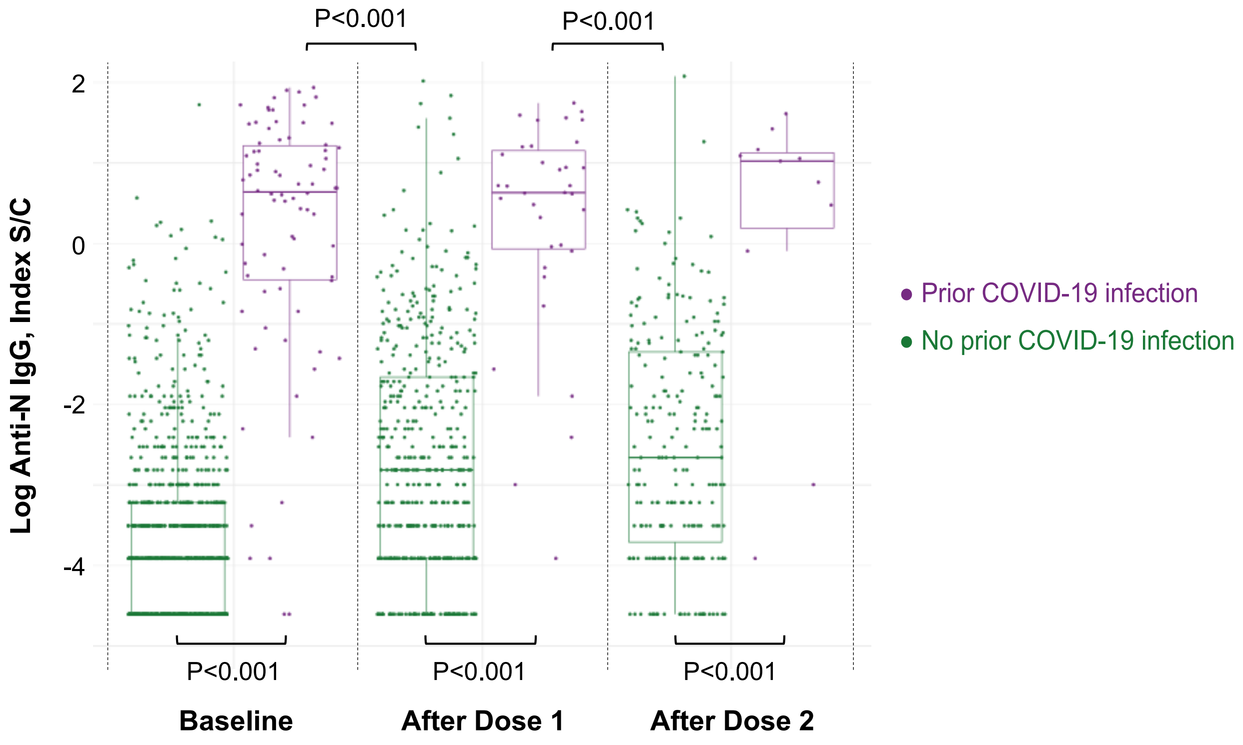

**Figure S2. Anti-Spike IgG Antibody Response to mRNA SARS-CoV-2 Vaccination in Persons With and Without Prior COVID-19 Infection: Values Above and Below 50 AU/mL.** Box plots display the median values with the interquartile range (lower and upper hinge) and ±1.5-fold the interquartile range from the first and third quartile (lower and upper whiskers).

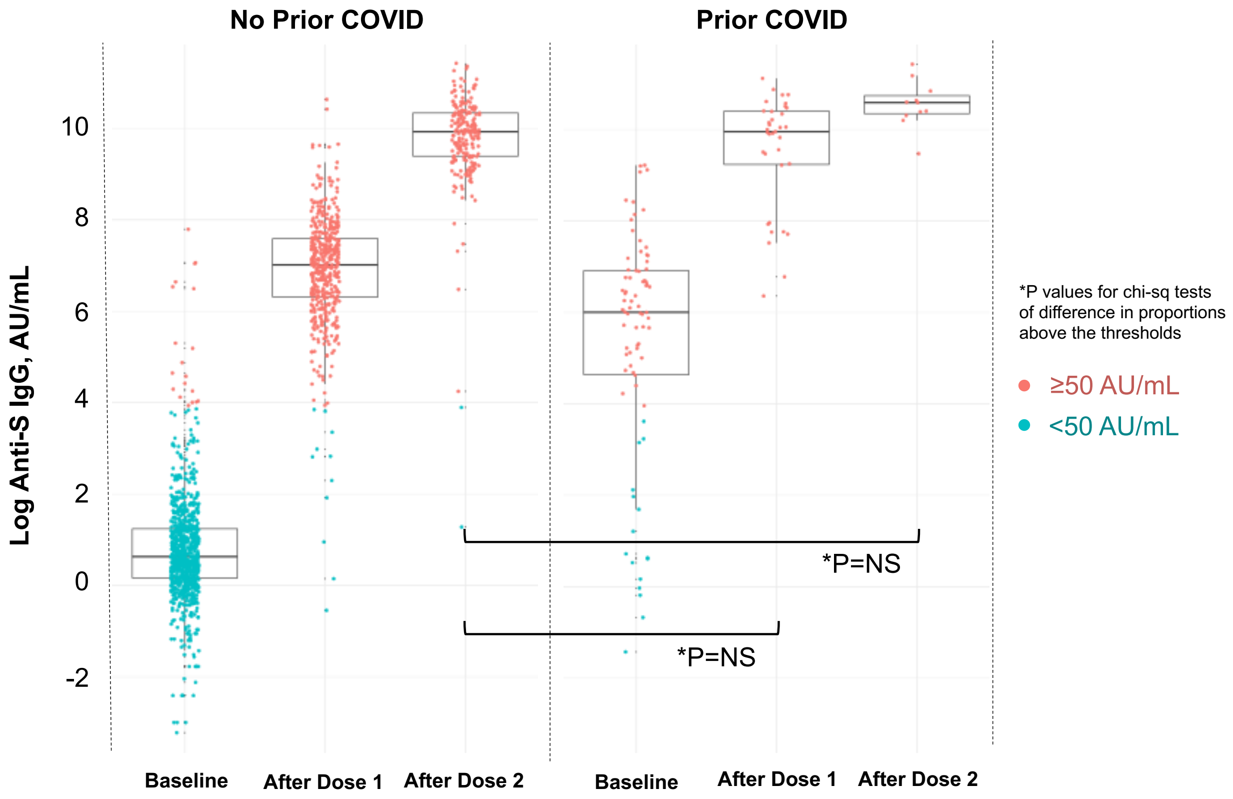

**Figure S3. Post-Vaccination Symptoms in Persons With and Without Prior COVID-19 Infection.**

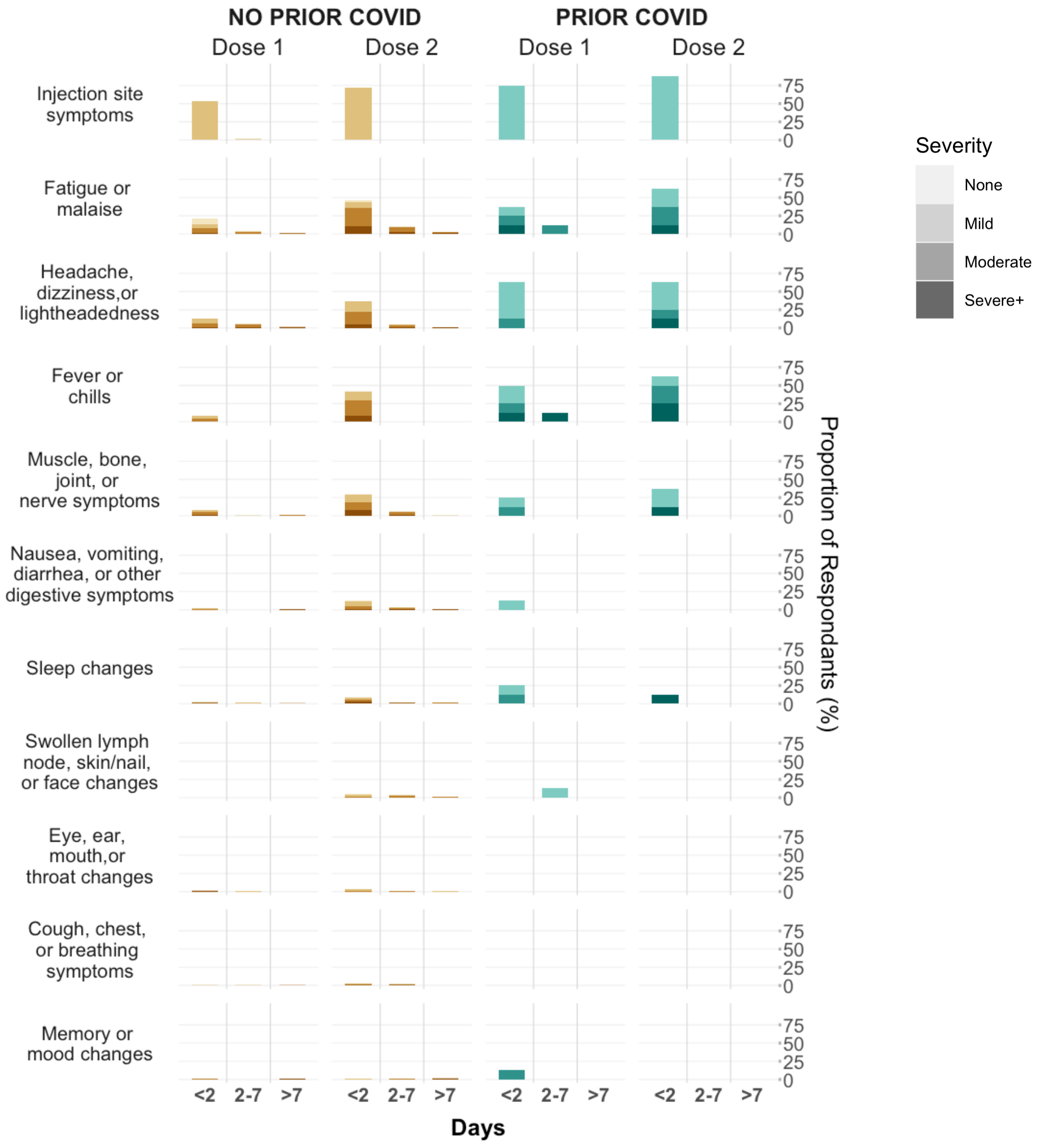

**Figure S4. Anti-Spike IgG Antibody Response to mRNA SARS-CoV-2 Vaccination in Persons With and Without Prior COVID-19 Infection: Values Above and Below 4160 AU/mL.** Box plots display the median values with the interquartile range (lower and upper hinge) and ±1.5-fold the interquartile range from the first and third quartile (lower and upper whiskers).

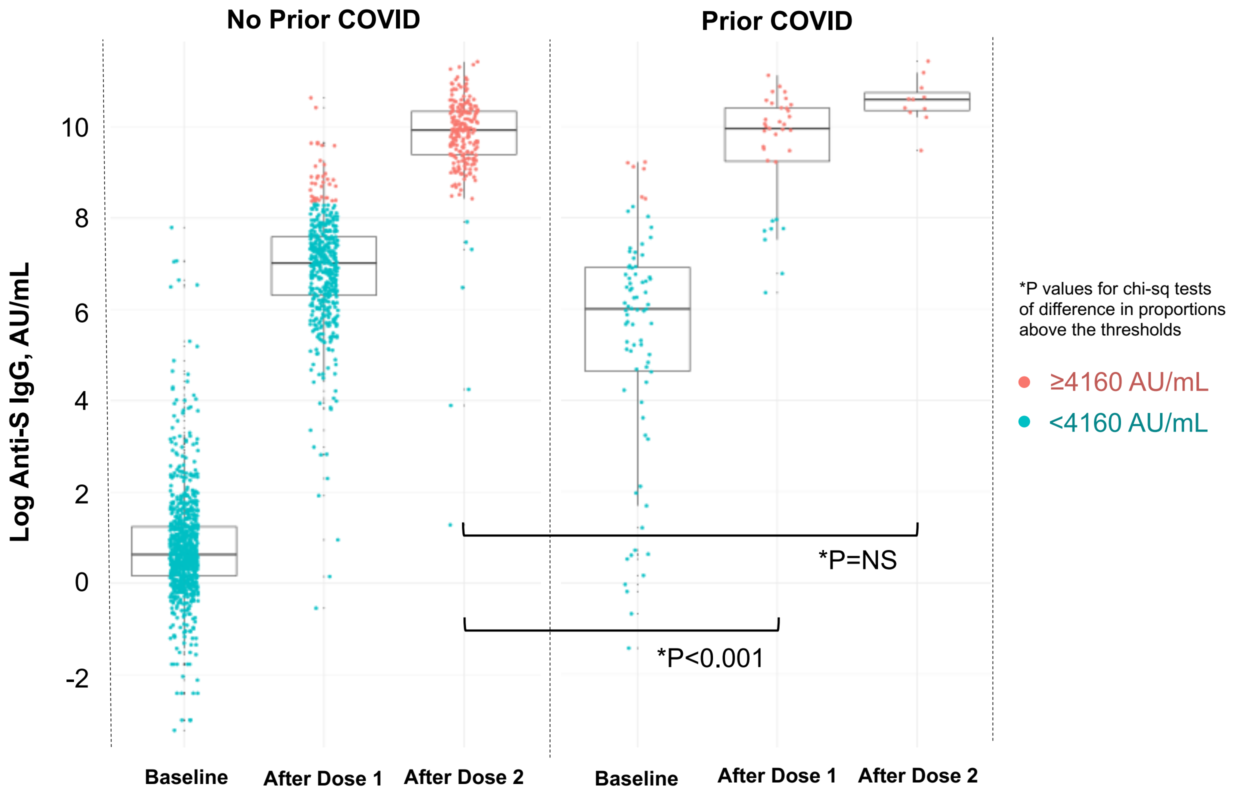
